# A systematic review of multi-variate time series approaches to extract predictive asthma biomarkers from routinely collected diary data

**DOI:** 10.1101/2024.01.31.24302056

**Authors:** Franz Aaron Clemeno, Matthew Richardson, Salman Siddiqui

## Abstract

**Objectives:** Longitudinal data is commonly acquired in asthma studies, to help assess asthma progression in patients, and to determine predictors of future outcomes, including asthma exacerbations and asthma control. Different methods exist for quantifying temporal behaviour in routinely collected diary variables to obtain meaningful predictive biomarkers of asthma outcomes. The aims of this systematic review were to evaluate the methods for extracting biomarkers from longitudinally collected diary data in asthma and investigate associations between the extracted measures and asthma patient reported outcomes (PROs).

**Setting:** A systematic review of MEDLINE, EMBASE, CINAHL and the Cochrane Library was conducted, using index terms relating to diary variables and asthma outcomes. Studies that focused on preschool children were excluded, to avoid confounding asthma with multi-factorial preschool wheeze. Study quality and risk of bias were assessed using the Transparent Reporting of a multivariable prediction model for Individual Prognosis Or Diagnosis (TRIPOD) and the Prediction model Risk Of Bias ASessment Tool (PROBAST), respectively.

**Participants:** Adults and/or children of school age (≥5 years old), with clinician-diagnosed asthma

**Primary outcomes:** Asthma PROs, namely asthma exacerbations, asthma control, asthma-related quality of life and asthma severity

**Results:** 24 full-text articles met the inclusion criteria and were included in the review. Generally, higher levels of variability in the diary variables were associated with poorer outcomes, especially increased asthma exacerbation risk, and poor asthma control. There was increasing interest in nonparametric methods to quantify complex behaviour of diary variables (6/24). TRIPOD and PROBAST highlighted a lack of consistent reporting of model performance measures and potential for model bias.

**Discussion:** Routinely collected diary variables aid in generating asthma assessment tools, including surrogate endpoints, for clinical trials, and predictive biomarkers of adverse outcomes, warranting monitoring through remote sensors. Studies consistently lacked robust reporting of model performance. Future research should utilise diary variable-derived biomarkers.

**Article Summary:** Strengths and limitations of this study

- This is the first systematic review that explores the different methods applied to time series of diary variables, namely peak flow, reliever use, symptom scores and awakenings.
- The scope of this review included multiple patient-reported outcomes, including asthma exacerbations, asthma control and asthma severity.

Only one reviewer was involved in screening the titles and abstracts for inclusion into the systematic review.

## Introduction

Asthma is a heterogeneous, chronic, inflammatory disorder of the airways [1] that currently affects millions of people worldwide [2]. It has been shown that there is wide variability between asthmatic patients in terms of the manifestations of asthma that they may experience [3], giving rise to different sub-types of asthma and potentially making it difficult to identify useful generalisable biomarkers to quantify disease activity.

Numerous outcome measures have been used to assess the state of asthma in patients. These include asthma exacerbation occurrence or risk, asthma control, asthma severity and asthma- and general health-related quality of life.

On top of these, numerous diary variables are often frequently captured longitudinally in studies to assist with asthma monitoring. Several approaches are available to analyse these time series, to extract biomarkers, to quantify the behaviour of diary variables, which could then be associated with and/or predictive of asthma outcomes. This includes simple measures, like mean and variance, non-parametric methods such as detrended fluctuation analysis (DFA) [4], which are well-suited for time series data, and machine learning approaches (ML), which can make use of rich, complex data. Extracting meaningful biomarkers from diary data is useful, since those variables are relatively easy and cheap to measure and record, as opposed to genomic or imaging data. Secondly, they can also be recorded with a high temporal granularity, thus giving a clear image in the longitudinal behaviour of the diary variables and disease progression.

Currently, no systematic review has been undertaken to assess the methodology applied to temporal diary variables, and the associations between the extracted measures and various patient reported outcomes (PROs) in asthma studies.

The objectives of this systematic review were to: 1) Review the methods used to extract biomarkers from longitudinally collected diary data, and 2) Report the associations between extracted measures and various asthma PROs.

## Methods

### Data Sources and Search Criteria

Four databases were searched in July 2023 namely Medical Literature Analysis and Retrieval System Online (MEDLINE), The Cochrane Library, Embase and Cumulative Index of Nursing and Allied Healthy Literature Plus (CINAHL). Only studies published from January 2000 and July 2023 were included in the review. The search strategy is provided as supplementary material.

Further details on methodology can be found in – **PROSPERO (CRD42021238910), Keywords and mesh terms included:** ‘asthma’, ‘peak expiratory flow’, ‘symptoms’, ‘reliever use’, ‘fractional exhaled nitric oxide’, ‘awakenings’, ‘exacerbations’, ‘control’, ‘severity’, and ‘quality of life’. The full search strategy used is outlined in Appendix 1 in the Supplementary Material.

### Inclusion and Exclusion Criteria

The review included studies of any type, such as: longitudinal, randomised controlled trials, open-label, retrospective analyses, etc.

The review only evaluated studies that used routinely collected data in their analyses with (I) a frequency of at least once daily over the course of (II) at least two weeks. Studies with a data collection frequency less than mentioned, or that had a shorter duration of follow-up were excluded from the review. This was to ensure that the diary variable data had sufficient temporal resolution and sampling duration to be able to realistically derive predictors of asthma outcomes.

The review only included studies whose participants were adults and/or children of school age (≥5 years old), with clinician-diagnosed asthma. Studies that focused on preschool children were excluded, to avoid confounding asthma with multi-factorial preschool wheeze.

### Review Process

Using the pre-defined inclusion and exclusion criteria, studies were initially selected from their title and abstracts. The full texts of these studies were then obtained, using either inter-library or British Library requests, were relevant. The full texts were again screened with the same inclusion criteria, where the reasons for exclusions were recorded. The reference lists of the included scrutinised to identify further relevant studies.

### Data Extraction

Data were extracted from the relevant studies, which included information regarding patient characteristics, diary variables of interest and their corresponding measures. Specifically, the study duration, sample size, the outcome variables and/or endpoints assessed, the diary variables used (from peak flow, night-time awakenings, reliever use and FeNO), the method/s of analysis and markers used in the analysis, and the summary of their findings.

### Study Quality Assessment

Assessment of risk of bias and quality of the included studies were conducted using PROBAST (Prediction model Risk Of Bias ASsessment Tool) [5] and the Transparent Reporting of a multivariable prediction model for individual prognosis or diagnosis (TRIPOD) checklist [6], respectively.

When the study quality was assessed, the percentage adherence of the studies to the different criteria of TRIPOD, as well as overall adherence of the studies to each criterion were recorded, to identify potential sources of bias.

## Results

### Studies identified

The literature search yielded 1,930 results across the four databases, of which 377 were excluded since they were duplicate studies. The remaining titles and abstracts were screened and narrowed down to 65 results for which full-text articles were sought. Using the pre-defined selection criteria, 48 of these results were excluded. Reasons for exclusion included conference abstract with no related full-text publication (n = 4), conference abstract with the full text included elsewhere in the literature search (n = 4), studies with data collection being too infrequent (i.e., less than at least once-daily over the course of at least two weeks) (n = 24), publication did not include any data (n = 7), studies where the variables were beyond the scope of the review (n = 8), or a publication was in a language such that translation services were not available (n=1). This left 17 studies for inclusion to the review. Their bibliographies were also searched for relevant papers, from which an additional 6 studies were identified for inclusion. Additionally, the bibliography of a systematic review that summarised the use of artificial intelligence (AI) in asthma [7] was searched for potentially relevant studies, and yielded one additional study. Overall, there were 24 studies included in the review. A Preferred Reporting Items for Systematic Reviews and Meta-Analyses (PRISMA) flow diagram is shown in **Figure 1. Table 1** summarises the 24 included studies.

**Figure 1.**
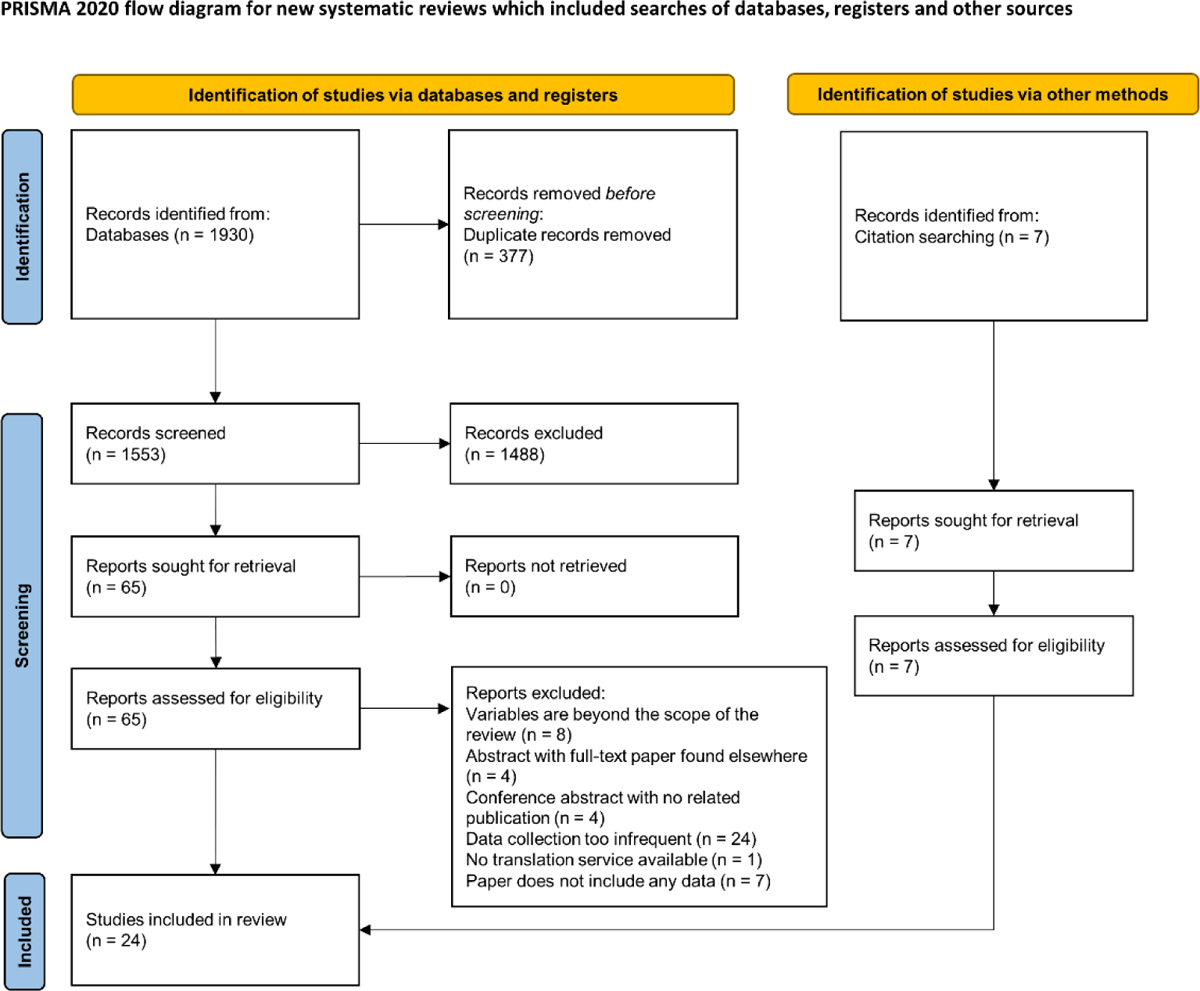
PRISMA flow chart illustrating study selection. (Modified from Page et al [47])

**Table 1:** Summary table of the included studies. Abbreviations used: PEF = peak expiratory flow; FeNO = fractional exhaled nitric oxide; DFA = detrended fluctuation analysis; AUC = area under curve; CV = coefficient of variation; AI = artificial intelligence; ROC = receiver operating characteristic; FEV1 = forced expiratory volume in 1 second; AQLQ = Asthma Quality of Life Questionnaire; ACT = Asthma Control Test; ICS = inhaled corticosteroid.

| Study | Modelled outcome variable/s | Duration | Sample Size | Diary variables used | Analysis method/s | Summary of findings |
| --- | --- | --- | --- | --- | --- | --- |
| Castner et al [8] | Asthma control | 6-8 weeks | 43 | Symptoms, awakenings | 3-day moving averages were calculated from diary data<br>Random effects models were used to identify significant predictors | 3-day moving averages of symptom scores were significantly predictive of asthma-specific awakenings and FEV1.<br>Wake counts were significantly associated with FEV1. |
| Covar et al [11] | Asthma exacerbations | 48 weeks | 285 | PEF, symptoms, reliever use | PEF values were summarised as seasonal averages in three-month blocks.<br>Regression modelling was used to investigate associations with occurrence of exacerbations.<br>Logistic regression models were used in the analysis. Univariate models were first built with each of the measures to narrow down the statistically significant covariates ($p < 0.05$) to include in the final model. | PEF expressed as the average drop in PEF over the entire season was significantly associated with the occurrence of exacerbations.<br>Diary variables evidently changed 12 to 3 days before an exacerbation, with a more apparent change within the last 2 days. Outcome measures returned to baseline within a maximum period of 10 days. |
| de Hond et al [34] | Asthma exacerbations | Development set: median 610 days<br>Validation set: median 417 days | Development set: 165<br>Validation set: 101 | PEF, reliever use, awakenings | Twice-daily measured PEF and reliever use, and nocturnal awakenings used as predictors. Various summary statistics (average, standard deviation, maximum and minimum) and time series transformations (first difference and lag) were also included in the models.<br>The performance of logistic regression was compared with one class SVM and XGBoost models to predict severe asthma exacerbation in a 2-day window. Their performance was compared to a proposed clinical rule (start oral corticosteroids treatment if PEF <60% personal best). | XGBoost achieved an AUC of 0.81, while logistic regression had an AUC of 0.88.<br>One class SVM had a lower sensitivity (0.34) than both XGBoost and logistic regression (0.6 and 0.73, respectively).<br>Both XGBoost and logistic regression achieved higher sensitivity than the clinical rule. |
| Finkelstein and Jeong [32] | Asthma exacerbations | Not stated | 26 | PEF, symptom score, reliever use, awakenings | Three classifiers were used: naïve Bayes, adaptive Bayesian network and support vector machines, using a 7-day window to predict exacerbation occurrence on day 8. | Using a 7-day window, the adaptive Bayesian network resulted in a perfect classification (sensitivity and specificity) |
|  |  |  |  |  |  | Shortening the time window resulted in worsened performance of the ML algorithms. |
| Frey et al [18] | Asthma exacerbations | 18 months | 80 | PEF | DFA was applied to the PEF time series to quantify its temporal behaviour, resulting in $\alpha$. The coefficient of variation (CV) was also calculated.<br>The conditional probability of experiencing a significant deterioration in airway obstruction was calculated. Calculation of the conditional probabilities was based on pre-specified threshold changes of PEF.<br>A nonlinear stochastic model of the fluctuations was introduced to assess the separate effects of the distribution and correlation of the PEF time series on the risk. | Lower $\alpha$ values generally reflect more severe airflow obstructions.<br>The risk of airway obstruction increases with decreasing $\alpha$ coupled with increased CV. |
| Fuhlbrigge et al [23] | Treatment response | 6/12 months | 17,415 | PEF, reliever use, symptom scores, awakenings | Threshold-based approach to develop event-based surrogate endpoint for severe exacerbations.<br>Various combinations of the four diary variables were evaluated to identify the optimal combination. Different combinations of the threshold and slope levels were also evaluated to select the most robust algorithm for CompEx.<br>The hazard ratio of CompEx was compared to that of severe exacerbations. | The final chosen algorithm was based on peak flow, reliever use and symptoms (PRS) (specific values in the original publication appendix Table ST1).<br>The proportion of patients with asthma exacerbation events increased 2.8 times, when censored at 3 months. Use of CompEx resulted in a net gain of power, with a 67% mean reduction in the number of patients required in a drug trial for severe exacerbations. |
| Greenberg et al [30] | Asthma exacerbations | 17 weeks | 1,114 | PEF, symptom scores, reliever use, awakenings | Disease activity was based on high and low cutoffs of daytime symptom score, awakenings, average rescue medication use, AQLQ-Activity domain, FEV1 and asthma attacks.<br>Stepwise, forward multiple regression analysis was used to determine which of the parameters to include in the weighted measure (ADAS-6). | Total β-agonist use/day, PEF and β-agonist use diurnal variability, and night-time awakenings contributed to the disease activity score, in a relatively balanced manner.<br>ADAS-6 discriminated between different levels of disease activity, as well as showing predictive validity for the risk of future asthma attacks. |
| Honkoop et al [25] | Asthma exacerbations | 1 year | 294 | PEF, symptom scores | Different action points (AP) were formed based on pre-specified thresholds for diary variables. Both univariate and multivariate action points were evaluated. Their performances were quantified using early detection days, sensitivity, specificity, Accuracy, AUC and NNT. | The optimal AP for early detection of asthma exacerbations was an increase in the composite symptoms score by greater than two standard deviations (from run-in) and a fall in PEF to <70% of their personal best occurring within a 1-week window. |
| Kaminsky et al [16] | Treatment response | 4-6 weeks run-in period<br>16-weeks treatment period | (1) 167<br>(2) 161<br>(3) 165 | PEF | The DFA coefficient $\alpha$ and coefficient of variation (CV) were calculated from PEF data during the run-in phase and the treatment phase. | An increase in $\alpha$ coupled with an increased CV is associated with treatment failure.<br>The pattern of alpha preceding treatment failure varied across the participants. |
| Khasha et al [31] | Asthma control | 9 months | 96 | PEF | Artificial intelligence (AI) model using ensemble learning, incorporating seven multiple base learners along with medical knowledge. Medical knowledge was incorporated through a rule-based classifier to classify the patient's asthma control level. | Morning and evening PEF were in the top three most important variables for asthma control level detection (alongside ACT score).<br>The proposed ensemble model had an accuracy of 0.943 for well-controlled zone, 0.894 for not well-controlled zone and 0.913 for very poorly controlled zone. |
| Kupczyk et al [12] | Asthma exacerbations | 1 year | Severe asthma 93<br>Mild to moderate asthma 76 | PEF, symptom score, reliever use | Looked at changes in diary variables at different periods pre-, during, and post-exacerbations.<br>ROC curves used to evaluate sensitivity and specificity of different threshold changes from baseline, to detect severe asthma exacerbations. | Regular monitoring of diary variables is able to detect severe exacerbations.<br>A 20% decrease in PEF resulted in sensitivity and specificity of 45% and 85% respectively.<br>For a 20% increase in daytime symptoms, the sensitivity and specificity were 46% and 84.9%. The combination of these two criteria resulted in the highest sensitivity and specificity: 65% and 94.9%. |
| Patel et al [13] | Asthma exacerbations, asthma control | 24 weeks | 147 in exacerbation analysis<br>142 in asthma control analysis | Reliever (salbutamol) use | Baseline reliever use metrics were used in the analysis.<br>Logistic regression was used to estimate the odds ratio for the association between the various metrics of salbutamol use and risk of severe asthma exacerbations, poor asthma control, and extreme salbutamol overuse. | Higher mean daily reliever, higher days of reliever, and higher maximal 24-hour use were associated with future severe exacerbations.<br>Higher mean daily use was associated with poor asthma control |
| Saito et al [10] | Asthma control, asthma severity | 2 weeks | 65 | PEF, FeNO | Diurnal variation, as well as other measures of daily and weekly variability was calculated for FeNO and PEF time series.<br>ROC curves were used to determine the detection of uncontrolled asthma.<br>Multivariate logistic regression analysis was used to identify predictors of uncontrolled asthma. | Diurnal variation of FeNO was able to discriminate between levels of asthma control, but diurnal variability of PEF was not able to.<br>Neither PEF or FeNO diurnal variability were able to discriminate between levels of asthma severity. |
| Spencer et al [26] | Asthma control | 1 year | 3,416 | PEF, reliever use, symptom scores, awakenings | Pre-specified thresholds of daytime symptom score, rescue beta2-agonist use, morning PEF, night-time awakening, asthma exacerbations, emergency visits, and treatment-related adverse events were used to determine the level of control each week (totally controlled (TC) or well-controlled (WC)).<br>Its relationship with the reference criteria (FEV1 and AQLQ) was tested using logistic regression models. | TC and WC asthma showed good discriminative properties when compared with % pred FEV1 and AQLQ at week 12 and change in % pred FEV1 from baseline to Week 52.<br>The composite asthma control measures have better discriminative properties compared to the individual asthma control status components alone. |
| Stern et al [22] | Asthma control, asthma exacerbations | 30 weeks | 41 | Symptoms, FeNO | DFA was applied to FeNO time series, resulting in the long-range scaling coefficient $\alpha$. Cross-correlation was calculated to quantify the linear correlation between FeNO and symptom scores.<br>Associations between the measures and outcomes of interest were evaluated using linear regression analysis. | Daily fluctuations in FENO values exhibited fractal-type long-range correlations. $\alpha$ values were significantly associated with baseline ICS use but were not associated with asthma control averaged over the whole period or in the last 12 weeks of the study.<br>Both $\alpha$ values and cross-correlation were able to distinguish between patients who experienced an exacerbation and those who did not. The cross-correlation between FeNO values and symptom scores was significantly higher in those subjects who had exacerbations. |
| Svensson et al [9] | Asthma exacerbations | 52 weeks | 502 | PEF, reliever use, symptom scores, awakenings | Diary variables were summarized as 4-day averages.<br>Stepwise covariate model selection was used to determine relevant predictors. | Diary variables showed trends 10-20 days prior to exacerbation events. PEF decreased and symptoms, reliever use, and awakenings increased. |
|  |  |  |  |  | An overall repeated time-to-event (RTTE) analysis was used to model repeated event data, which incorporates baseline and time-varying covariates, as well as treatment exposure and baseline hazard. | Symptom score and rescue medication were significant in the model as time-varying covariates.<br>PEF was significant in the forward selection but not in the backward selection. |
| Thamrin et al 2009 [19] | Treatment response | 6 months for each treatment period | 66 | PEF, symptom score | DFA was applied to PEF time series, using only 300 data points for each treatment period, which corresponds to the first 150 days of each treatment period. The resulting value is denoted $\alpha$.<br>Regression models used to examine associations between predictors of interest and the clinical outcomes. | $\alpha$ calculated from the placebo period was significantly associated with treatment response to salmeterol, where higher values suggested a decrease in symptom days. |
| Thamrin et al 2010 [14] | Asthma control | 2 weeks pre-withdrawal period<br>6 weeks or until loss of control post-withdrawal, whichever came first | 83 | PEF | Variability measures were calculated both pre- and post-withdrawal, including the coefficient of variation of PEF (CV).<br>Cox regression was used to explore associations between time to loss of asthma control (LOC) and the variability measures. ROC curves were used to assess the utility of each of the measures for predicting LOC. | An increase in CV in any of the three periods were significant predictors of LOC in the separate models. Autocorrelation was not a significant predictor of LOC, even when the trend was removed.<br>The larger the increase in CV within 2 weeks of ICS withdrawal, the sooner the patient will experience LOC. |
| Thamrin et al 2011a [21] | Asthma exacerbations | Study A: 52 weeks<br>Study B: 72 weeks | Study A: 77<br>Study B: 58 | PEF | The conditional probability is calculated from simulated PEF time series, using DFA to quantify the correlation properties.<br>Calculation of the conditional probabilities was based on pre-specified threshold changes of PEF.<br>Logistic regression was used to examine associations between conditional probabilities and PEF events and asthma exacerbations. | The conditional probability was related to actual decreases in PEF. A 10% increase in the probability was associated with the risk of having a future exacerbation. |
| Thamrin et al 2011b [20] | Asthma control, asthma exacerbations, asthma severity | 6 months | Study 1: 132<br>Study 2: 159 | PEF | DFA was performed on twice-daily PEF to derive $\alpha$.<br>Significant predictors were identified using multinomial or binary logistic regression, | $\alpha$ values were able to discriminate between levels of asthma control, where lower values were found in patients with uncontrolled asthma. |
|  |  |  |  |  | depending on the outcome. | $\alpha$ values did not differ significantly between severity groups.<br>Patients with exacerbations had significantly higher $\alpha$ values than those who did not. |
| Van der Valk et al [17] | Asthma exacerbations | 30 weeks | 27 | Reliever use, FeNO | Daily FeNO and symptom scores were taken from 3-week blocks. Fluctuation and correlation metrics used were coefficient of variation, the slope of daily FeNO, cross-correlation and autocorrelation.<br>Parameters were evaluated as predictors for exacerbations using logistic regression. | The following FeNO parameters were associated with moderate exacerbation risk:<br>- CV (14-10 days before)<br>- Slope (14-4 days before)<br>- Cross-correlation (14-10 days before)<br>- Autocorrelation (21-10 days before)<br>There was marked variability in FeNO but no clear rise preceding the onset of severe exacerbations. There was no clear trend of symptoms relative to severe exacerbations. |
| Van Vliet et al [27] | Asthma control | 1 year | 78 | Symptoms | Asthma control by home monitoring was determined based on data 1 week before the clinical visit and was calculated based on GINA criteria for asthma control (threshold-based approach).<br>Agreement between the two methods was analyzed by the linear Cohen kappa coefficient. | There was low agreement between the 2 instruments to distinguish the 3 levels of asthma control. |
| Wu et al [15] | Asthma exacerbations | 4 years | 1,019 | Symptoms | Symptom scores were aggregated in four-month blocks. Pre-specified thresholds were used to classify patients as to whether they experienced persistent symptoms. Generalized estimating equation (GEE) models were used to study predictors of persistent symptoms, as well as severe exacerbations. | Symptom category is associated with severe exacerbations even when adjusting for demographic, pulmonary and biologic measures.<br>Predictors of both persistent symptoms and severe exacerbations include ICS treatment (budesonide), FEV1/FVC ratio, and PC20. |
| Zhang et al [33] | Asthma exacerbations | Mean 362 days | 2,010 | PEF, reliever use, symptom scores, awakenings | Four machine learning models were trained and tested, namely logistic regression, naïve Bayes, decision trees and perceptrons. | The best model used logistic regression with input variables derived from principal components analysis. |
|  |  |  |  |  |  | The model had an area under the receiver operating characteristic curve of 0.85, with a sensitivity of 90% and specificity of 83% for detecting severe asthma exacerbations up to three days before its occurrence. |

### Diary Variable Usage

The usage of diary variables in the studies are shown in **Figure 2a**. From the included studies, peak expiratory flow (PEF) was the most used diary variable, with 18 studies including it in their analyses. Conversely, night-time awakenings were the least used, with only 8 studies using it in their analyses. Symptom scores and short-acting bronchodilator reliever use were used in 14 and 11 of the studies, respectively. Nine of the included studies used only one diary variable in their analyses, and 6 used all of four. Fractional Exhaled Nitric Oxide (FeNO) was included in 4 of the studies, where it was used as a diary variable in 3 of them.

**Figure 2.**
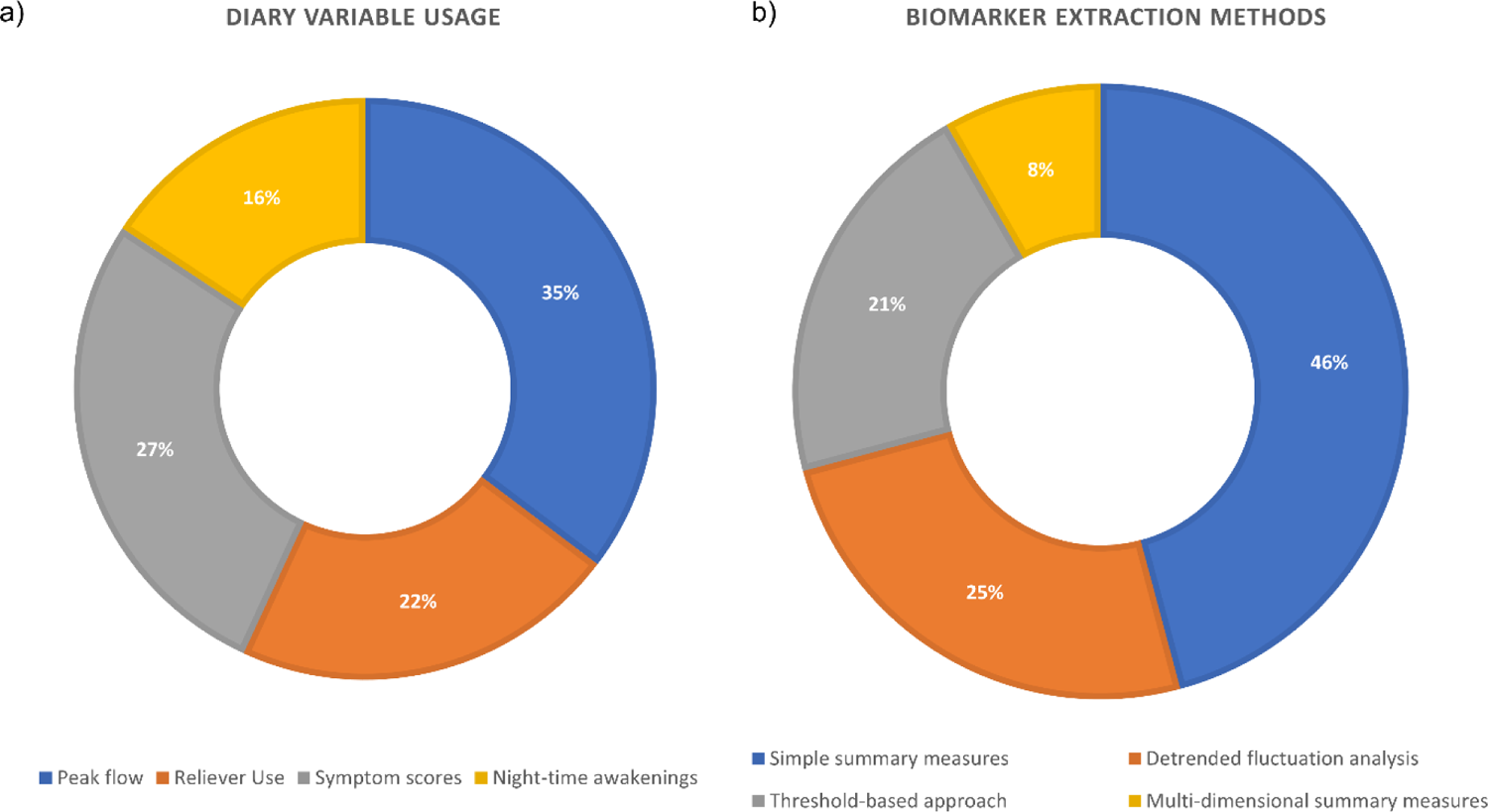
Breakdowns of the included studies by (a) diary variable usage; (b) biomarker extraction methods.

### Biomarker extraction methods

The methods used to extract biomarkers from diary variables are summarised in **Figure 2b**. Several studies used simple summary measures to quantify the behaviour of the diary variables throughout the observation periods. These include moving averages [8, 9], diurnal variability [10], seasonal/periodic averages [11–15], coefficient of variation [14, 16, 17], and autocorrelation [17]. Overall, these studies showed that increased variability in the diary variables is associated with more adverse outcomes, namely exacerbation risk, loss of asthma control and treatment failure to inhaled steroids. Additionally, higher levels of symptom scores, reliever use, and night-time awakenings were also associated with increased exacerbation risk or occurrence, and poorer asthma control. Conversely, decreases in PEF were associated with increased exacerbation risk or occurrence. The cross-correlation between daily FeNO and symptom scores were also associated with moderate exacerbation risk, where stronger correlations between the two variables was associated with increased risk [17].

A non-parametric approach, detrended fluctuation analysis (DFA) was used in 6 of the studies, 5 of which applied it to time series of PEF recordings, and 1 to time series of FeNO measurements. DFA quantifies the strength of long-range correlations in the time series through the resulting long-range scaling coefficient, denoted α. Four of the PEF studies [16, 18–20] used DFA to extract biomarkers as potential predictors of asthma PROs and the other [21] solely used DFA to simulate additional PEF time series. These studies show that α is related to asthma PROS, specifically the risk of exacerbations and airway obstruction. Some studies report that a lower α is indicative of increased risk of airway obstruction [18], but some found that higher values may be indicative risk of treatment failure to inhaled steroids, when coupled with an increase in the coefficient of variation of PEF [16]. Lower α values were also found in patients with uncontrolled asthma, but α values did not differ significantly between asthma severity groups [20]. The DFA coefficient α from PEF during the placebo period was also shown to predict treatment response to salmeterol, but notably, not salbutamol, where higher values of α during the placebo period was associated with improved treatment response. DFA was also applied to time series of FeNO data, and one study found significantly increased α in patients who had experienced an exacerbation [22].

### Threshold-based approaches

Several studies used pre-specified threshold changes in diary variables over pre-specified windows of time to develop markers, and surrogate or early endpoints of asthma PROs.

Fuhlbrigge et al. aimed to develop an intermediate endpoint for asthma exacerbations using diary variables [23]. The endpoint was defined based on prespecified threshold changes or worsening (slope) greater than some prespecified magnitude, over at least 2 or 5 days, respectively. These thresholds were amalgamated with the ATS/ERS definition of asthma exacerbations, defined by oral steroid treatment utilisation [3], yielding a composite score. The final endpoint, denoted CompEx only included PEF, reliever use and symptom scores (CompEx-PRS). CompEx-PRS identified an increased exacerbation event frequency by 2.8-fold and whilst preserving treatment effect sizes observed on exacerbations.

Kupczyk et al. also utilised multiple diary variables and aimed to find a proxy for exacerbations [24]. A 20% decrease in PEF or a 20% increase in day symptoms on 2 consecutive days was able to detect severe exacerbations with a sensitivity of 65% and a specificity of 95%, where combining the two improved the overall predictive performance.

Honkoop et al. aimed to validate optimal action points of PEF and symptoms to aid with early detection of exacerbations [25]. The optimal combination (PEF and symptoms) action point comprised an increase of more than two standard deviations of the symptom score from the run-in mean, and a decrease of PEF to <70% of their personal best. This action point detected exacerbations 1.4 days before their occurrence with 80.5% sensitivity and 98.3% specificity.

Wu et al. used simple thresholds to aggregate daily diary card scores into a symptom score for each 4-month block and evaluated its associations with severe exacerbation occurrence [15]. Symptom scores were associated with severe exacerbations, where patients with more blocks of persistent symptoms being more likely to experience more exacerbations during the 4-year study.

Spencer et al. validated a composite measure of asthma control [26]. The measure was comprised of daytime symptom score, rescue beta2-agonist use, morning PEF, night-time awakening, asthma exacerbations, emergency visits, and treatment-related adverse events and used simple pre-specified thresholds to determine asthma control level. The resulting measure showed good discriminative ability of other measures of asthma control, both cross-sectionally and longitudinally.

Van Vliet et al. compared two methods for assessing asthma control, namely prospective symptom and lung function monitoring versus retrospective recall using the Asthma Control Questionnaire (ACQ) [27]. Prospective assessment of asthma control was measured using daily symptom questionnaires and FEV1 values and using thresholds to classify the level of asthma control on a weekly basis, based on GINA control criteria [28]. Conversely, retrospective assessment of asthma control was conducted using the ACQ during the routine clinic visits. There was low concordance between the two methods, but it seems that prospective monitoring provides a more realistic image of patient health, potentially since it minimises recall bias for retrospective recall.

Frey et al. [18] and Thamrin et al. [29] both used threshold changes of PEF to calculate the conditional probability of an airway obstruction, defined as PEF <80% (moderate) or PEF <60% (severe) of the age- and height-predicted normal values occurs within a certain time period [28] given a patient’s current PEF value, denoted π. As previously mentioned, Frey found that airway obstruction risk was associated with increased variability and loss of deterministic behaviour of PEF. Thamrin found that π was associated with actual occurrences of airway obstructions. Additionally, π was shown to be associated with future exacerbation risk, where an increase in this probability was associated with an increase in the odds ratio of having a future exacerbation.

Greenberg et al. used a threshold-based approach to develop a composite score, named ADAS-6, comprised of rescue beta-agonist use (daily use and diurnal variability), PEF diurnal variability, and night-time awakenings, as well as FEV1 % predicted and AQLQ (symptom domain score), to determine the level of disease activity in patients [30]. The authors defined disease activity based on high and/or low cut-offs for the following variables: daytime symptom score, night-time awakenings, average rescue beta-agonist use, AQLQ score (activity domain), FEV1 % predicted, and asthma attacks. ADAS-6 was discriminative of disease activity and demonstrated content and convergent validity. The study found that each of the 6 included variables contributed to the regression models in a relatively, balanced manner, looking at their standardized coefficients.

### Machine Learning and Artificial Intelligence Approaches

Of the studies in the review, 4 analysed data, including the diary variables of interest using machine learning (ML) and AI. These studies aimed to build predictive models.

Several algorithms were used, namely ensemble learning, Naïve Bayes, support vector machines (SVM), adaptive Bayesian networks, XGBoost, one class SVM, logistic regression, decision trees and perceptrons. ML models demonstrated good predictive performance in the studies.

Khasha et al. used an ensemble model, which combined numerous disease-related variables and and medical knowledge to detect asthma control level, which was determined using a rule-based classifier derived from the physicians’ knowledge [31]. The resulting classifier had a good performance, with an accuracy of over 91%. Interestingly, among the large number of variables used in the algorithm, morning, and evening PEF, along with ACT score, as a measure of daily symptoms, were the most important features.

Finkelstein and Jeong utilised diary data collected through telemonitoring and evaluated three ML methods to build a predictive model for early prediction of asthma exacerbations [32]. The authors used a naïve Bayesian classifier, adaptive Bayesian network and SVM, of which the adaptive Bayesian network performed best, resulting in a perfect classification in terms of sensitivity, specificity, and accuracy when a 7-day window was used to predict an exacerbation on the eighth day.

Zhang et al. was also interested in predicting exacerbation occurrence using daily diary data, but whether an exacerbation occurred on the same day or up to three days in the future [33]. The authors evaluated the performance of several ML methods, namely, logistic regression, decision tree, naïve Bayes classifier, and perceptron algorithms. The best performing model was logistic regression applied to data processed using principal component analysis (PCA), and achieved ROC = 0.85, sensitivity = 90%, specificity = 83% for detecting severe asthma exacerbations.

De Hond et al. [34] developed and compared predictive models for the early detection of severe asthma exacerbations, using a 2-day prediction horizon. The authors compared the performances of two ML models (XGBoost and one class SVM), a logistic regression model and a simple asthma action plan. The logistic regression model (AUC = 0.88) outperformed the XGBoost model (AUC = 0.81), as well as the one class SVM model. Notably, both the XGBoost and logistic regression models reached higher discriminative performance compared to the simple clinical rule.

With the extracted biomarkers, regression was the most utilised class of methods for assessing their associations with the PROs. Of the included studies, 15 used a regression method in their analyses. These include many different classes of models, including linear, multinomial, random effects, Cox, etc. A few studies used more complex regression models, such as repeated time-to-event analysis [9] and generalised estimating equations [15].

### Risk of Bias and Quality Assessment

None of the studies showed full compliance to the TRIPOD reporting criteria. The percentage adherence of reporting to the TRIPOD criteria is shown in **Figure 3**, broken down by study (**Figure 3a**) and by criterion (**Figure 3b**). Lack of adherence was prominent in the reporting of key elements of the study setting, participant flow, and model performance measures, where of the 24 studies, only 9 (38%), 6 (25%) and 12 (52%) studies reported these, respectively. Of the separate criteria, only 1 of the studies reported the key study dates, specifically the start and end of accrual. Some of these can be attributed to the fact that 16 of the studies were retrospective, and so simply referred to the original publications for that information.

**Figure 3.**
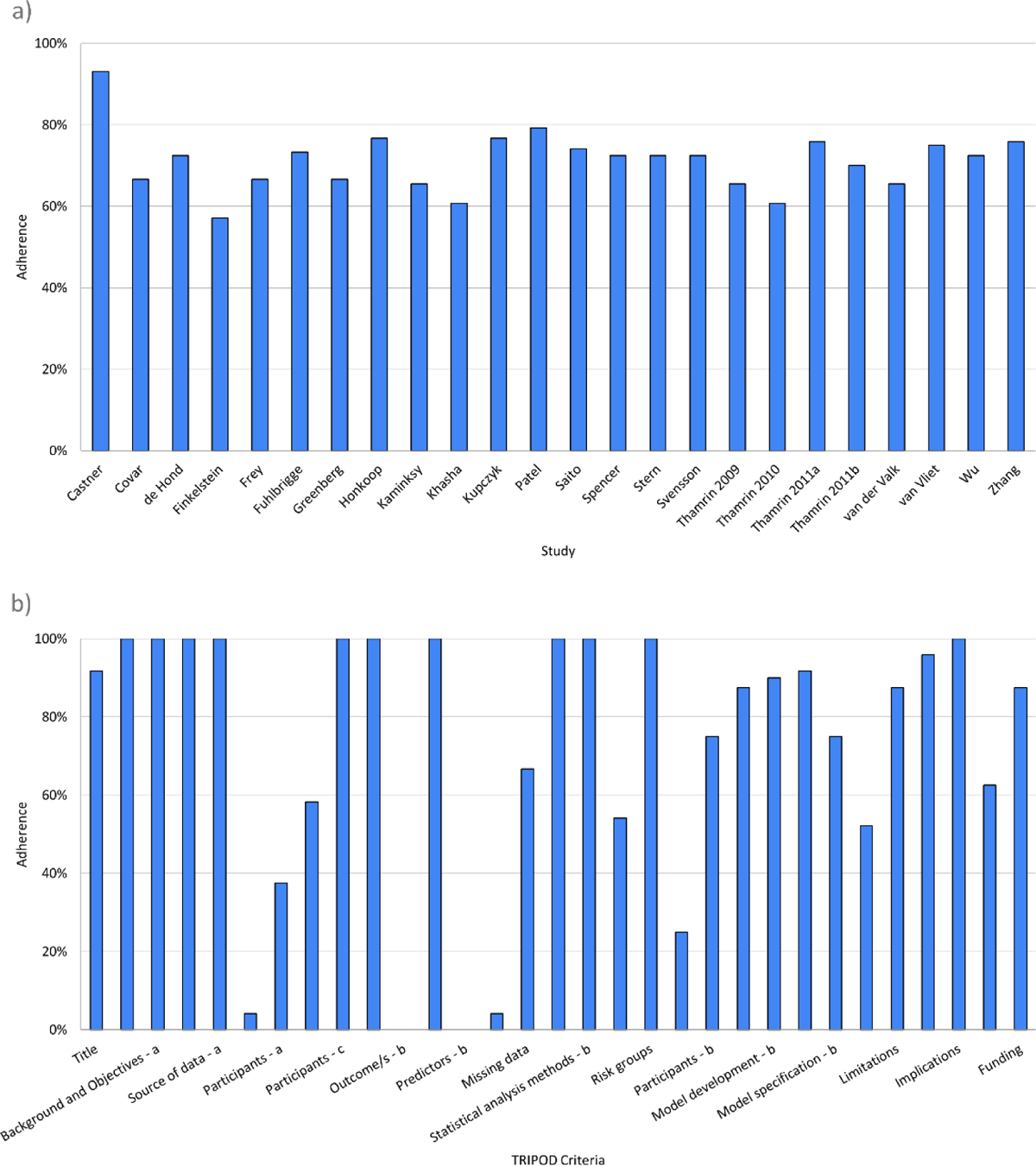
Percentage compliance to TRIPOD checklist of the included studies, broken down by: (a) study; (b) separate criteria.

Assessment of risk of bias using PROBAST can be found in **Figure 4**. One prominent source of bias among the included studies were in Domain 4: Analysis. Many studies did not appropriately report model performance measures, specifically regarding model overfitting, underfitting and optimism, where only 7 studies reported the appropriate measures. This finding is confirmatory of those observed when looking at study adherence to TRIPOD. None of the included studies were deemed to have a high risk of bias overall.

**Figure 4.**
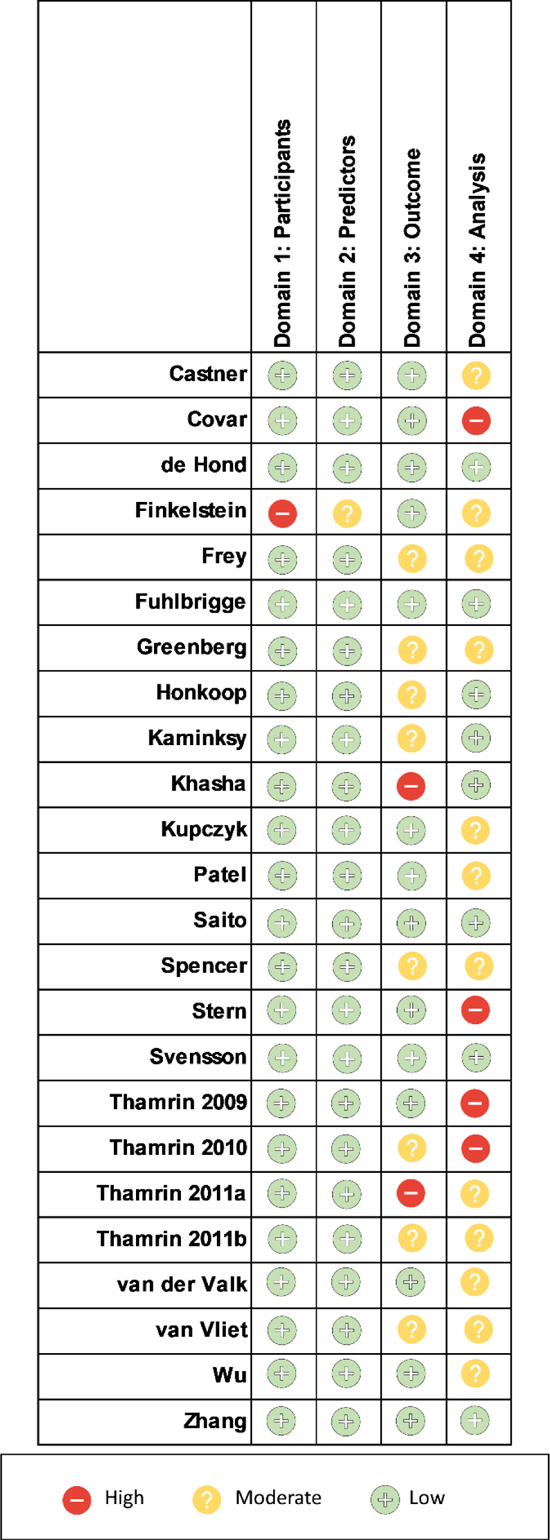
Risk of bias assessment using the PROBAST signalling questions.

Another source of potential bias that is not addressed in the PROBAST tool is recording bias, which comes from studies not using electronic or automated method to record diary variables. 10 of the 24 studies (42%) recorded patient data on paper diaries.

## Discussion

This systematic review provided an overview of methodologies applied to routinely collected diary variables in adult asthma, namely, PEF, FeNO, reliever use, night-time awakenings, and symptoms to extract biomarkers of PROs, namely asthma exacerbations, asthma control and asthma severity, of which 24 studies were identified. The heterogeneity of the studies, in terms of outcome variables and extracted measures meant that conducting a meta-analysis was not possible.

Of the diary variables of interest, PEF was the most used, potentially because it is both an index of airflow obstruction and the only continuous variable of those included and can hence demonstrate more complex behaviour. Conversely, night-time awakenings were the least used, possibly due to them usually being included as a binary variable. Reliever use and symptom scores were often included as discrete scores, which can display some level of complex temporal behaviour, but not to the extent of PEF. Despite the differences in the type of variables, they demonstrated balanced contributions to asthma disease activity [30].

Interestingly, only a few studies recorded FeNO as a diary variable. This is despite FeNO being a biomarker of airway inflammation and exhibiting long-range temporal correlations. Additionally, its joint behaviour with symptom scores, quantified by cross-correlation, was significantly associated with asthma exacerbations occurrence [22]. Further research should investigate complex temporal behaviour of FeNO as potential predictors of asthma PROs.

Few studies used ML and AI models to build predictive models, even though routinely collected diary variables can provide numerous data points per patient. The studies that opted to use these methods [31–33] built predictive models which showed decent performance. A systematic review of AI in asthma found a growing interest in the use of such methods, but there is currently a lack of research in the context of asthma treatment, especially to biologics [7]. One study [34] however demonstrated that ML models do not always outperform classic statistical models, such as logistic regression, and that researchers should consider whether the complexity of the data warrants the use of complex ML methods. Future studies can use ML models to utilize the complex temporal behaviour of the data to predict PROs, alongside other data streams simultaneously, such as demographic, genomic, etc. data.

The systematic review has demonstrated that biomarkers that quantify the temporal behaviour of diary variables are associated with asthma PROs, namely asthma exacerbations, asthma control and treatment response, or failure. Generally, higher levels of variability in the diary variables were associated with increased risk of asthma exacerbations, and uncontrolled asthma. Threshold-based approaches were the most common among the included studies for extracting biomarkers from the diary variables. These methods were useful in identifying and quantifying simple temporal changes in the diary variables over a short pre-specified period. Threshold approaches resulted in markers that were able to detect the occurrence of asthma exacerbations early.

Temporal variability was also quantified using a range of simple summary measures, as well as novel nonparametric methods, such as DFA. DFA first introduced by Peng and was initially applied to time series of DNA nucleotides [4] and later to time series of heart rate [35]. DFA quantifies fractal scaling properties of time series, as well as detecting and quantifying the long-range correlations of nonstationary time series, into one quantitative parameter α. DFA has been applied to numerous physiological time series, such as to detect irregularities in the heart rate of patients with severe congestive heart failure [36], as well as to differentiate between healthy and diabetic patients, the fluctuations in their plasma glucose [37]. DFA has also been used to identify long-term self-similarity in the daily PEF of COPD patients, and that α was significantly associated with exacerbation frequency [38]. This systematic review identified an increasing use of DFA, which quantifies more complex long-range scaling temporal behaviour of the diary variables, compared to simple summary measures and threshold-based approaches. DFA has shown promise in generating biomarkers, which are associated with PROs, including asthma exacerbations and treatment response. Despite being mainly applied to time series of PEF data, application to FeNO data has shown promise and should be further explored in future studies.

Future studies can also employ alternative methods to quantify temporal behaviour. One example is entropy [39], which is a method that has been applied to biological signals in patients with asthma [40]. A systematic review of the applications of entropy in asthma found that the entropy values of signals such as respiratory sound and airway resistance, and respiratory inter-breath interval, are strong potential novel indices of asthma progression and asthma severity, respectively [41].

For identifying and quantifying associations between the extracted markers and PROs, regression methods were the most common. Regression models have model coefficients and results which are often easy to interpret for researchers and clinicians, and many classes of regression models are available to handle different data types.

The review demonstrates that inclusion of more than one diary variable in analyses showed utility, as different measures capture different aspects of asthma. The temporal relationship between FeNO and symptoms, quantified using cross-correlation was shown to be a potential predictor of exacerbations [22]. Greenberg used multiple diary variables to develop a score that measured disease activity and was related to asthma attack occurrence [30]. Interestingly, the diary variables contributed equally to the score and removing diurnal variability worsened its performance, especially in small sample sizes. Future studies should consider temporal relationships between diary variables as potential predictors of asthma PROs.

The included studies demonstrated partial adherence to TRIPOD criteria, specifically with reporting model performance measures. A systematic revew of prediction models for future asthma exacerbations similarly report a lack of robust validation analyses to demonstrate generalisability of results [42]. Future studies should improve reporting adherence to TRIPOD, to enable qualification of markers as asthma endpoints.

Based on the PROBAST tool, none of the studies had an overall high risk of bias. One significant source of bias was a lack of sufficient reporting in terms of the model performances in concordance with the TRIPOD analysis. While this doesn’t affect the models’ performances, it helps readers determine the generalisability of the developed models for readers. Another source of potential bias was from manual collection of diary variables. This introduces potential measurement and recall bias, especially if patients were expected to collect and record this data themselves, rather than by a physician.

Another potential source of bias is the consistency and consideration of the data collection times [43]. Asthma demonstrates significant diurnal rhythmicity, where symptoms generally worsen overnight or early in the morning [44]. Conversely, night-time PEF and FEV1 are reduced compared to day-time values [45]. Additionally, studies have shown that there is within-day variability in airway inflammation, measured using FeNO, even in stable asthmatics [46]. These suggest that timing of the diary variable collection is important, and that consistent timings in the collection is vital in minimising the effect of within-day variability due to circadian rhythms, which would potentially mask fluctuations caused by disease.

One limitation of this review is that only one reviewer (FC) was responsible for the screening of the studies. This could have introduced potential bias in the selection of studies.

The included studies show that routinely collected diary variables demonstrate clinical utility in two domains: 1. The generation of asthma assessment tools, such as surrogate markers or early endpoints, which can aid researchers and clinicians to design shorts and more powerful clinical trials; 2. The discovery and/or generation of biomarkers or models, which are predictive of adverse outcomes in asthma, including asthma exacerbations. This warrants the use of remote sensors and electronic diaries for daily monitoring of asthma patients, to help identify those who are at high risk of an asthma exacerbation. These are summarised in Supplemental **Figure 1**.

In summary, this review highlights the importance of quantifying the longitudinal, often complex behaviour of daily-recorded diary variables and their utility in developing biomarkers that are predictive of asthma outcomes, namely asthma exacerbations, asthma control, asthma severity and asthma-related quality of life. Consequently, future research should expand nonparametric methods that can be used to quantify this behaviour, alongside standardising both the capture of diary data, in view of the growing number of digital devices and m-health technology used to acquire diary variables in asthma. Future studies should adhere to robust multivariable model reporting standards, specifically in terms of model performance and validation, as to allow for qualification of the biomarkers in the context of interventional studies.

## Data Availability

All data produced in the present work are contained in the manuscript.

## Acknowledgments

We thank Pip Divall (Glenfield Hospital Library, Glenfield Hospital, Leicester, United Kingdom) and Selina Lock (University of Leicester, Leicester, United Kingdom) for their invaluable contributions to the systematic literature search.

## References

1. National Asthma, E., A.E. Prevention Program, and P. Prevention, Expert Panel Report 3 (EPR-3): Guidelines for the Diagnosis and Management of Asthma– Summary Report 2007. Journal of allergy and clinical immunology, 2007. 120(5): p. S94–S138.

2. Morris, M.J. Asthma. 2020 [cited 2021 25/10/2021]; Available from: https://emedicine.medscape.com/article/296301-overview#a6.

3. Reddel, H.K., et al., An Official American Thoracic Society/European Respiratory Society Statement: Asthma Control and Exacerbations: Standardizing Endpoints for Clinical Asthma Trials and Clinical Practice. American journal of respiratory and critical care medicine, 2009. 180(1): p. 59–99.

4. Peng, C.K., et al., *Mosaic organization of DNA nucleotides.* Physical review. E, Statistical physics, plasmas, fluids, and related interdisciplinary topics, 1994. 49(2): p. 1685–1689.

5. Wolff, R.F., et al., PROBAST: A Tool to Assess the Risk of Bias and Applicability of Prediction Model Studies. Annals of Internal Medicine, 2019. 170(1): p. 51–58.

6. Collins, G., et al., Transparent reporting of a multivariable prediction model for individual prognosis or diagnosis (TRIPOD): The TRIPOD statement.

7. Exarchos, K.P., et al., Artificial Intelligence techniques in Asthma:A systematic review and critical appraisal of the existing literature. The European respiratory journal, 2020. 56(3): p. 2000521.

8. Castner, J., et al., Prediction model development of women’s daily asthma control using fitness tracker sleep disruption. Heart & Lung, 2020. 49(5): p. 548–555.

9. Svensson, R.J., et al., Population repeated time-to-event analysis of exacerbations in asthma patients: A novel approach for predicting asthma exacerbations based on biomarkers, spirometry, and diaries/questionnaires. CPT: pharmacometrics & systems pharmacology, 2021. 10(10): p. 1221–1235.

10. Saito, J., et al., Domiciliary diurnal variation of fractional exhaled nitric oxide (FeNO) to monitor asthma control. European Respiratory Journal, 2013. 42.

11. Covar, R.A., et al., Factors associated with asthma exacerbations during a long-term clinical trial of controller medications in children. Journal of allergy and clinical immunology, 2008. 122(4): p. 741–747.e4.

12. Kupczyk, M., et al., Detection of exacerbations in asthma based on electronic diary data: results from the 1-year prospective BIOAIR study. Thorax, 2013. 68(7): p. 611.

13. Patel, M., et al., Metrics of salbutamol use as predictors of future adverse outcomes in asthma. Clinical and experimental allergy, 2013. 43(10): p. 1144–1151.

14. Thamrin, C., et al., Variability of lung function predicts loss of asthma control following withdrawal of inhaled corticosteroid treatment. Thorax, 2010. 65(5): p. 403–408.

15. Wu, A.C., et al., Predictors of symptoms are different from predictors of severe exacerbations from asthma in children. CHEST, 2011. 140(1): p. 100–107.

16. Kaminsky, D.A., et al., Fluctuation analysis of peak expiratory flow and its association with treatment failure in asthma. American Journal of Respiratory and Critical Care Medicine, 2017. 195(8): p. 993–999.

17. Van Der Valk, R.J.P., et al., Daily exhaled nitric oxide measurements and asthma exacerbations in children. American Journal of Respiratory and Critical Care Medicine, 2010. 181(1).

18. Frey, U., et al., Risk of severe asthma episodes predicted from fluctuation analysis of airway function. Nature, 2005. 438(7068): p. 667–70.

19. Thamrin, C., et al., Fluctuation analysis of lung function as a predictor of long-term response to beta2-agonists. The European respiratory journal, 2009. 33(3): p. 486–493.

20. Thamrin, C., et al., Associations between fluctuations in lung function and asthma control in two populations with differing asthma severity. Thorax, 2011. 66(12): p. 1036–1042.

21. Thamrin, C., et al., Predicting future risk of asthma exacerbations using individual conditional probabilities. Journal of Allergy and Clinical Immunology, 2011. 127(6): p. 1494–1502.e3.

22. Stern, G., et al., Fluctuation phenotyping based on daily fraction of exhaled nitric oxide values in asthmatic children. The Journal of allergy and clinical immunology, 2011. 128(2): p. 293–300.

23. Fuhlbrigge, A.L., et al., A novel endpoint for exacerbations in asthma to accelerate clinical development: a post-hoc analysis of randomised controlled trials. The Lancet. Respiratory medicine, 2017. 5(7): p. 577–590.

24. Kupczyk, M., et al., Detection of exacerbations in asthma based on electronic diary data: results from the 1-year prospective BIOAIR study. Thorax, 2013. 68(7): p. 611–8.

25. Honkoop, P.J., et al., Early detection of asthma exacerbations by using action points in self-management plans. The European respiratory journal, 2013. 41(1): p. 53–59.

26. Spencer, S., et al., Validation of a guideline-based composite outcome assessment tool for asthma control. Respiratory research, 2007. 8: p. 26.

27. van Vliet, D., et al., Electronic monitoring of symptoms and lung function to assess asthma control in children. Annals of Allergy, Asthma & Immunology, 2014. 113(3): p. 257–262.e1.

28. Global Initiative for Asthma, Global Strategy for Asthma Management and Prevention. 2012: www.ginasthma.org.

29. Thamrin, C.P., et al., Predicting future risk of asthma exacerbations using individual conditional probabilities. Journal of allergy and clinical immunology, 2011. 127(6): p. 1494–1502.e3.

30. Greenberg, S.M.D., et al., The Asthma Disease Activity Score: A discriminating, responsive measure predicts future asthma attacks. Journal of allergy and clinical immunology, 2012. 130(5): p. 1071–1077.e10.

31. Khasha, R., M.M. Sepehri, and S.A. Mahdaviani, An ensemble learning method for asthma control level detection with leveraging medical knowledge-based classifier and supervised learning. Journal of medical systems, 2019. 43(6): p. 1–15.

32. Finkelstein, J. and I.c. Jeong, Machine learning approaches to personalize early prediction of asthma exacerbations. Annals of the New York Academy of Sciences, 2017. 1387(1): p. 153–165.

33. Zhang, O., L.L. Minku, and S. Gonem, Detecting asthma exacerbations using daily home monitoring and machine learning. The Journal of asthma: official journal of the Association for the Care of Asthma, 2020: p. 1–10.

34. de Hond, A.A.H., et al., Machine learning did not beat logistic regression in time series prediction for severe asthma exacerbations. Scientific reports, 2022. 12(1): p. 20363.

35. Peng, C.-K., et al., Quantification of scaling exponents and crossover phenomena in nonstationary heartbeat time series. Chaos An Interdisciplinary Journal of Nonlinear Science, 1995. 5: p. 82.

36. Carvalho, T.D., et al., Fractal correlation property of heart rate variability in chronic obstructive pulmonary disease. International journal of chronic obstructive pulmonary disease, 2011. 6(1): p. 23–28.

37. Ogata, H., et al., The lack of long-range negative correlations in glucose dynamics is associated with worse glucose control in patients with diabetes mellitus. Metabolism, clinical and experimental, 2012. 61(7): p. 1041–1050.

38. Donaldson, G.C., et al., Detrended fluctuation analysis of peak expiratory flow and exacerbation frequency in COPD. The European respiratory journal, 2012. 40(5): p. 1123–1129.

39. Richman, J.S. and J.R. Moorman, Physiological time-series analysis using approximate entropy and sample entropy. American Journal of Physiology-Heart and Circulatory Physiology, 2000. 278(6): p. H2039–H2049.

40. Gonem, S., et al., Airway impedance entropy and exacerbations in severe asthma. The European respiratory journal, 2012. 40(5): p. 1156–63.

41. Sun, S., et al., *Entropy Change of Biological Dynamics in Asthmatic Patients and Its Diagnostic Value in Individualized Treatment: A Systematic Review.* Entropy (Basel, Switzerland), 2018. 20(6): p. 402.

42. Bridge, J., J.D. Blakey, and L.J. Bonnett, A systematic review of methodology used in the development of prediction models for future asthma exacerbation. BMC medical research methodology, 2020. 20(1): p. 22–22.

43. Wang, R., et al., Asthma diagnosis: into the fourth dimension. Thorax, 2021. 76(6): p. 624–631.

44. Turner-Warwick, M., Nocturnal asthma: A study in general practice. Journal of the Royal College of General Practitioners, 1989. 39(323): p. 239–243.

45. Martin, R.J., L.C. Cicutto, and R.D. Ballard, Factors related to the nocturnal worsening of asthma. The American review of respiratory disease, 1990. 141(1): p. 33–38.

46. Wilkinson, M., et al., Circadian rhythm of exhaled biomarkers in health and asthma. The European respiratory journal, 2019. 54(4): p. 1901068.

47. Page, M.J., et al., The PRISMA 2020 statement: an updated guideline for reporting systematic reviews. BMJ, 2021. 372: p. n71.

