## Supplementary material for "A systematic review of multi-variate time series approaches to extract predictive asthma biomarkers from routinely collected diary data": Online Supplement

Norfolk Place

London W2 1PG

**Authors’ affiliations**

^1^Institute for Lung Health, NIHR Leicester Biomedical Research Centre, University of Leicester, Leicester, United Kingdom

^2^National Heart and Lung Institute, Imperial College, London, United Kingdom

**Appendix 1**

The search strategy that was used for all four databases (EMBASE, MEDLINE, CINAHL, The Cochrane Library) is outlined below:

1.	exp Asthma/

2.	asthma*.mp.

3.	1 or 2

4.	"diary variable".mp.

5.	"diary data".mp.

6.	"lung function".mp.

7.	Peak Expiratory Flow Rate/

8.	"peak expiratory flow".mp.

9.	"reliever use".mp.

10.	"rescue medication".mp.

11.	"inhaler use".mp.

12.	awakening*.mp.

13.	symptom score*.mp.

14.	"airway inflammation".mp.

15.	Biomarkers/

16.	fractional exhaled nitric oxide.mp.

17.	Symptom Flare Up/

18.	symptom*.mp.

19.	4 or 5

20.	7 or 8

21.	9 or 10 or 11

22.	13 or 17 or 18

23.	14 or 15 or 16

24.	6 or 12 or 19 or 20 or 21 or 22 or 23

25.	Patient Reported Outcome Measures/

26.	patient reported outcome*.mp.

27.	exacerbation*.mp.

28.	asthma attack*.mp.

29.	Airway Obstruction/

30.	"airway obstruction".mp.

31.	"airway deterioration".mp.

32.	"asthma episode".mp.

33.	"Quality of Life"/

34.	"quality of life".mp.

35.	"Asthma control".mp.

36.	"asthma severity".mp.

37.	25 or 26

38.	27 or 28 or 29 or 30 or 31 or 32

39.	33 or 34

40.	35 or 36 or 37 or 38 or 39

41.	3 and 24 and 40

42.	limit 41 to yr="2000 -Current"

43.	limit 42 to english language

**Figure Legends**

**Supplemental Figure 1.** Clinical utility of longitudinally collected diary variables.

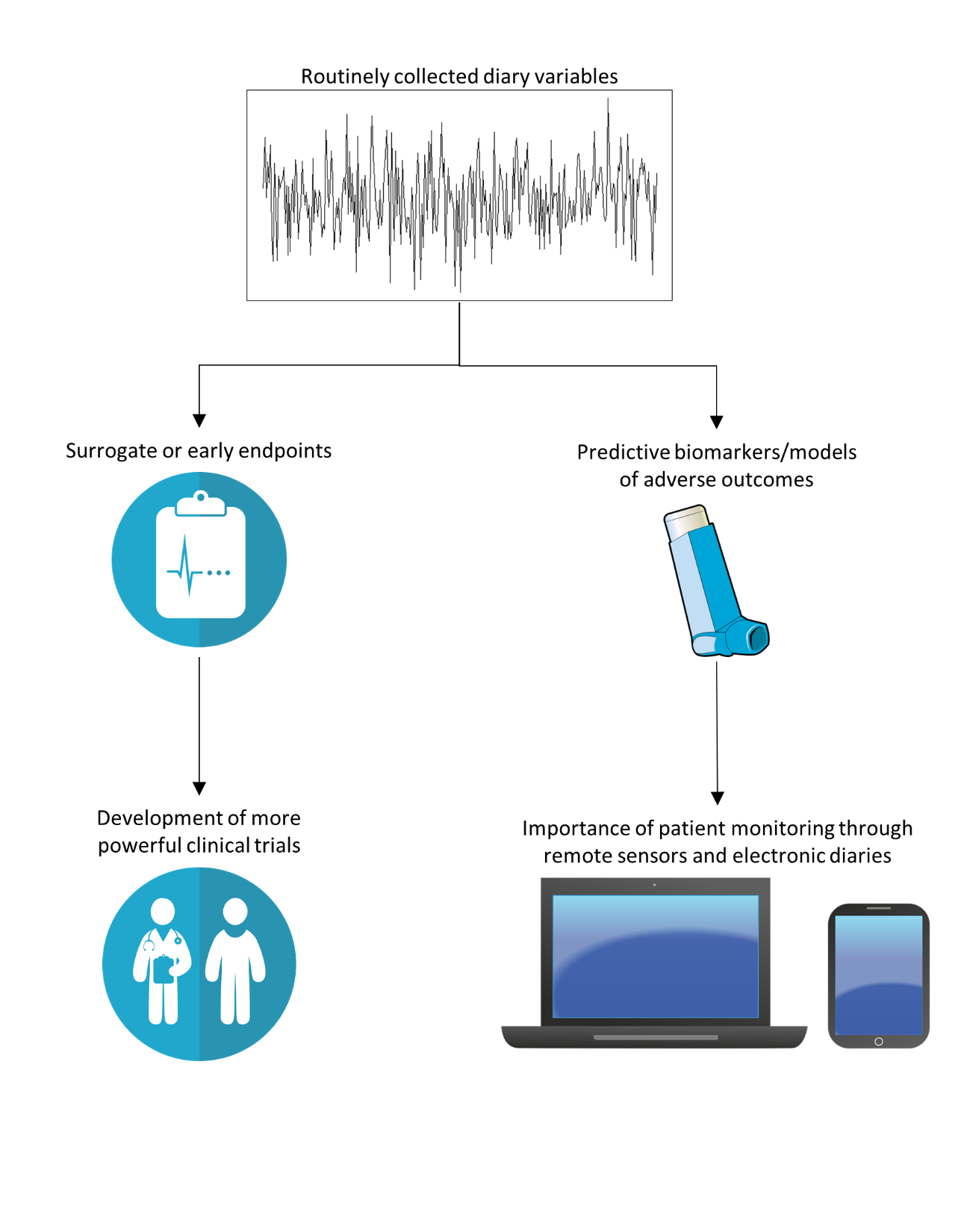
